# Health impacts of expanding different health workforce cadres under a limited budget in Malawi

**DOI:** 10.1101/2025.09.13.25335695

**Authors:** Bingling She, Sakshi Mohan, Rachel E. Murray-Waston, Sangeeta Bhatia, Martin Chalkley, Tim Colbourn, Joseph H. Collins, Emilia Connolly, Eva Janoušková, Dominic Nkhoma, Paul Revill, Asif U. Tamuri, Pakwanja D. Twea, Tara D. Mangal, Joseph Mfutso-Bengo, Timothy B. Hallett, Margherita Molaro

## Abstract

Like many others, Malawi’s health care system faces significant health workforce shortages largely due to budget constraints that limit training, recruitment and retention of staff. A crucial question is how to best allocate a limited budget to expand different health care workers (HCW) cadres so that the potential health gains are maximised, which is more important now than ever considering recent withdraw and reduction in donor funding. This research aims to provide a practical answer to this question. We designed a range of budget allocation scenarios for HCW expansion across different cadres, and used the “all diseases – whole healthcare system” *Thanzi La Onse* (TLO) model to estimate the resulting population health outcomes. We find that, indeed, how to allocate additional resources for HRH across different cadres is an important determinant of potential health impact. Putting all resources into increasing staffing in a single cadre is not the most effective use of the resources, even if that cadre has the most limited availability currently. Similarly, allocating new resources in a manner that mirrors the current distribution of spending does not generate the greatest possible gains. Instead, an allocation that uplifts staffing for all cadres, according to extra time and cost required to meet the arising healthcare needs, gives the greatest benefits. We conclude that in the context of complex interplay between demography, epidemiology, treatment scope and effectiveness, and health resource constraints in the health care system, human resources for health (HRH) bottlenecks in achieving health gains are multifactorial and a balanced mix of cadres and skills is required for future HRH expansion. As such, health system models such as the TLO that capture this interplay can make potential contributions to strengthening HRH planning.

**What is already known on this topic:** To date, very few studies have quantitatively analysed the potential future health impact of HRH expansion in the context of evolving healthcare needs for a whole population. One existing study shows that investing in HRH expansion has the potential to achieve better health outcomes for Malawians, assuming a uniform expansion of multiple cadres (1).

**What this study adds:** Under a detailed individual-based simulation model capturing the wide range of healthcare needs and the interdependency between cadres for delivering care, we have shown that there is not a simple HRH bottleneck driving future health outcomes and health gains could be achieved by allocating additional resources to expand cadres in a carefully balanced way.

**How this study might affect research, practice or policy:** This study answers the hard question of how to use the limited funding for HRH expansion to achieve greatest health gains in Malawi; and demonstrates the essential use of system-wide modelling to support decision-making in complex health systems.

## Introduction

Health systems in Malawi and many other low- and middle-income countries are challenged with shortages of human resources for health (HRH) and underfunding, in conjunction with increasing health care needs from growing populations (2-8). In such resource-constrained settings, the decision to expand HRH requires careful considerations of how limited funds are allocated between in health care worker (HCW) cadres.

This is a critical but hard question to answer in practice, considering the evolving health care needs and health burdens in a complex health care system that is undergoing demographic, epidemiologic and economic changes, a detailed representation of the cadres required for delivering that care, and an understanding of the consequences that follow if the care is revived or if it is not. Whereas many studies have estimated the degree of HCW shortages based on recorded, targeted or simulated service levels (2, 3, 5, 9-13), few studies have examined the health consequences of such shortages or aimed to find the optimal resource allocation for HRH expansion by cadres. A linear constrained optimisation method has been used to investigate the marginal health impacts of investing in individual health worker cadres in Uganda and Malawi, assuming static constraints and epidemiological profile (4, 14). However, as these analyses were not attached to epidemiological models, it is hard to estimate the health consequences of alternative resource allocations in a way that takes into account the interlocking epidemiological dynamics. System dynamics modelling (SDM) has also been used to inform health workforce planning. In this approach, macro-level health system behaviours, such as changes or movements of resources/services over time, are incorporated and the health workforce needs and health outcomes are normally driven by the changes of health care services that are strategically designed (15, 16). In such an approach, though, the micro-level interactions between in health system resources for delivering care and patients needing care cannot be considered, making it difficult to explore how detailed changes of HCW cadres would affect health service provision and population health outcomes that are dependent on such interactions.

The *Thanzi La Onse* model (17-19), an individual-based simulation model for Malawi health care system, captures demographic and epidemiologic changes and interactions between individuals and the health care system; thereby providing a new means to examine the health consequences of HRH expansion addressing these gaps. Using this model, we have already quantified the potential health gains that could result from a uniform expansion of HCW cadres (6). This study now turns to explore how additional resources could be allocated across cadres for future expansions to maximise health gains in a complex, resource-constrained system.

## Methods

### The Thanzi La Onse model

The Thanzi La Onse (TLO) model is an individual-based simulation model of health-care service delivery in the public sector in Malawi. It models the health care needs of the population and how health care system resources including HRH, consumables, facilities and equipment are used to meet these needs, and simulates health service provision and health burden in terms of disability-adjusted life years (DALYs) of the population. Capabilities of HRH (i.e., total working time for delivering patient-facing/centred services) and consumables (i.e., the current probability of availability of drugs and medical supplies), health system function in terms of diagnostic accuracy, referral practice and HCW competence, healthcare seeking behaviour, and service prioritisation policy are modelled constraints or parameters that impact the health service delivery and health outcome. Further details of this model and parameters can be found elsewhere (1, 17, 20-22).

### HRH expansion scenarios

We model the expansion of the HRH in TLO model by allocating an incremental budget among HCW cadres every year over the period between 2025 and 2034 (inclusive), and examine the health impacts of different allocation scenarios. We consider 5 groups of cadres - Clinical (C), Nursing and Midwifery (NM), Pharmacy (P), Disease Control and Surveillance Assistant or Health Surveillance Assistant (D), and Other (O) that includes Dental health, Laboratory, Mental health, Nutrition and Radiography cadres. These groups include all main cadres in the current workforce (excluding administrative staff) as detailed in Appendix A.1.1.

Following Molaro *et al*. (1), we assume a yearly HRH budget growth rate *R* = 4.2%, i.e., the incremental budget for HRH expansion for the next year is 4.2% of the total HRH cost of the previous year. We also assume the incremental budget and HRH cost here refer to staffing salaries only.

Each budget allocation scenario specifies what proportion of that incremental budget goes toward funding each of the 5 cadres, considering the differing salaries across cadres (See salary details and the scenario illustration diagram and in Appendices A.1.1 and A.1.2). We have designed 33 allocation scenarios (as listed in Table 1) that can be grouped as below.

**Table 1.**
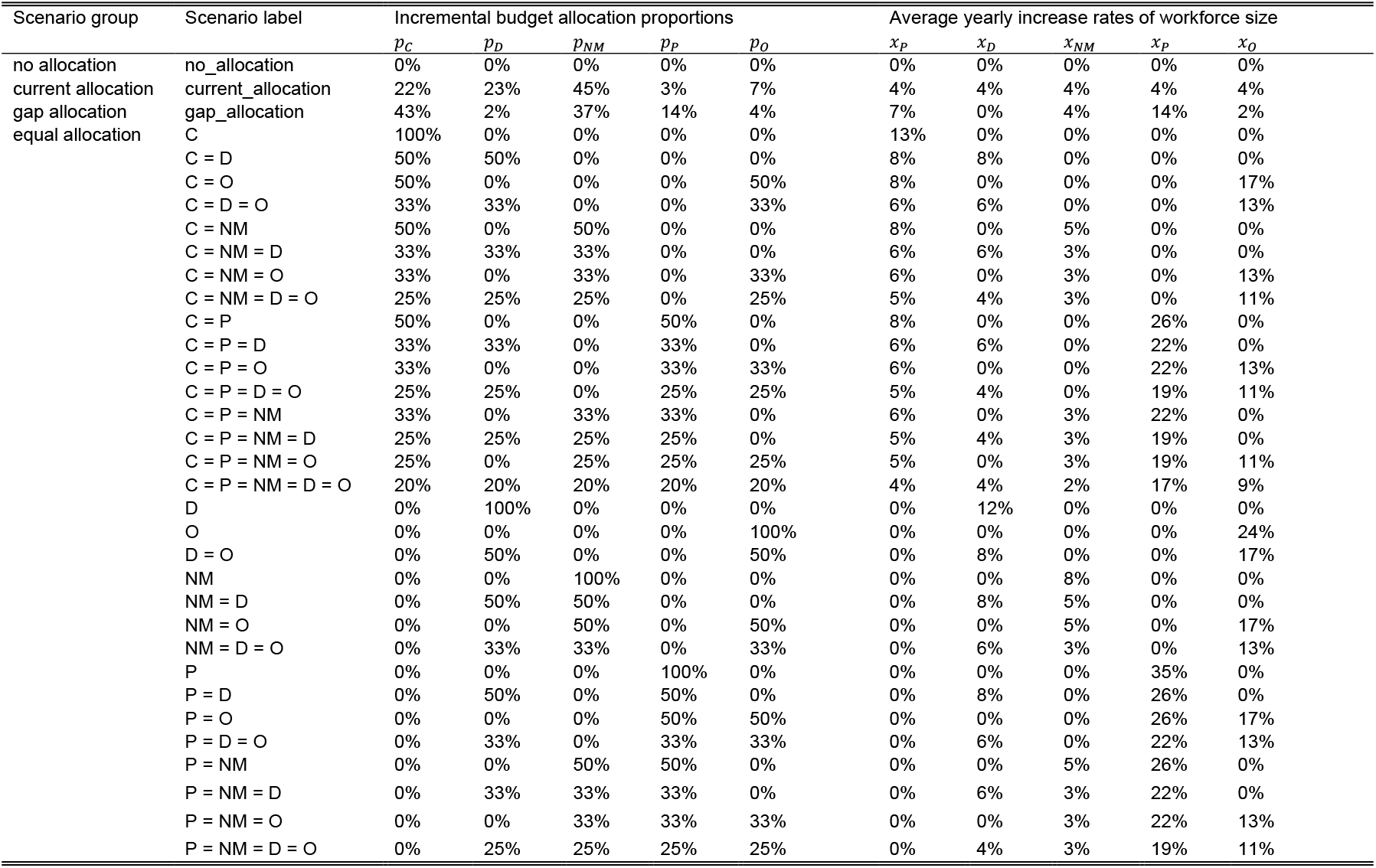
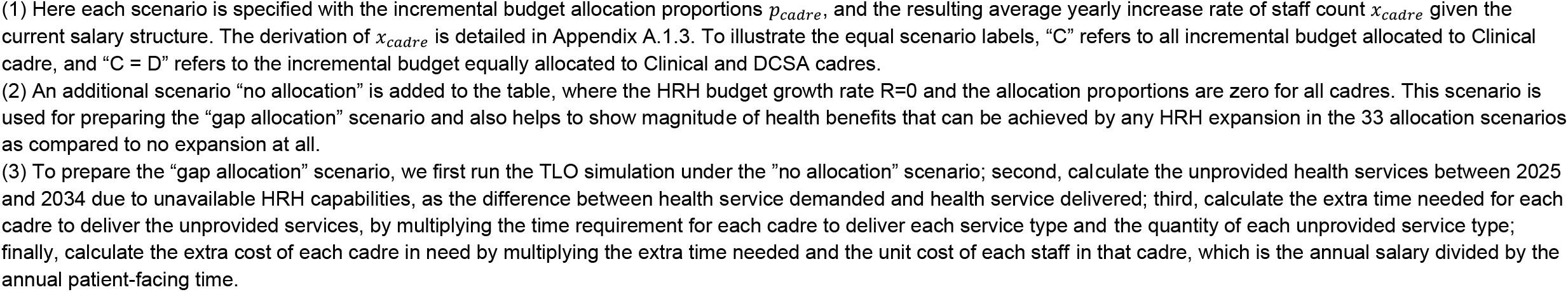
The incremental budget allocation scenarios.

| Scenario group | Scenario label | Incremental budget allocation proportions |  |  |  |  | Average yearly increase rates of workforce size |  |  |  |  |
| --- | --- | --- | --- | --- | --- | --- | --- | --- | --- | --- | --- |
| | | $p_C$ | $p_D$ | $p_{NM}$ | $p_P$ | $p_O$ | $x_P$ | $x_D$ | $x_{NM}$ | $x_P$ | $x_O$ |
| no allocation | no_allocation | 0% | 0% | 0% | 0% | 0% | 0% | 0% | 0% | 0% | 0% |
| current allocation | current_allocation | 22% | 23% | 45% | 3% | 7% | 4% | 4% | 4% | 4% | 4% |
| gap allocation | gap_allocation | 43% | 2% | 37% | 14% | 4% | 7% | 0% | 4% | 14% | 2% |
| equal allocation | C | 100% | 0% | 0% | 0% | 0% | 13% | 0% | 0% | 0% | 0% |
|  | C = D | 50% | 50% | 0% | 0% | 0% | 8% | 8% | 0% | 0% | 0% |
|  | C = O | 50% | 0% | 0% | 0% | 50% | 8% | 0% | 0% | 0% | 17% |
|  | C = D = O | 33% | 33% | 0% | 0% | 33% | 6% | 6% | 0% | 0% | 13% |
|  | C = NM | 50% | 0% | 50% | 0% | 0% | 8% | 0% | 5% | 0% | 0% |
|  | C = NM = D | 33% | 33% | 33% | 0% | 0% | 6% | 6% | 3% | 0% | 0% |
|  | C = NM = O | 33% | 0% | 33% | 0% | 33% | 6% | 0% | 3% | 0% | 13% |
|  | C = NM = D = O | 25% | 25% | 25% | 0% | 25% | 5% | 4% | 3% | 0% | 11% |
|  | C = P | 50% | 0% | 0% | 50% | 0% | 8% | 0% | 0% | 26% | 0% |
|  | C = P = D | 33% | 33% | 0% | 33% | 0% | 6% | 6% | 0% | 22% | 0% |
|  | C = P = O | 33% | 0% | 0% | 33% | 33% | 6% | 0% | 0% | 22% | 13% |
|  | C = P = D = O | 25% | 25% | 0% | 25% | 25% | 5% | 4% | 0% | 19% | 11% |
|  | C = P = NM | 33% | 0% | 33% | 33% | 0% | 6% | 0% | 3% | 22% | 0% |
|  | C = P = NM = D | 25% | 25% | 25% | 25% | 0% | 5% | 4% | 3% | 19% | 0% |
|  | C = P = NM = O | 25% | 0% | 25% | 25% | 25% | 5% | 0% | 3% | 19% | 11% |
|  | C = P = NM = D = O | 20% | 20% | 20% | 20% | 20% | 4% | 4% | 2% | 17% | 9% |
|  | D | 0% | 100% | 0% | 0% | 0% | 0% | 12% | 0% | 0% | 0% |
|  | O | 0% | 0% | 0% | 0% | 100% | 0% | 0% | 0% | 0% | 24% |
|  | D = O | 0% | 50% | 0% | 0% | 50% | 0% | 8% | 0% | 0% | 17% |
|  | NM | 0% | 0% | 100% | 0% | 0% | 0% | 0% | 8% | 0% | 0% |
|  | NM = D | 0% | 50% | 50% | 0% | 0% | 0% | 8% | 5% | 0% | 0% |
|  | NM = O | 0% | 0% | 50% | 0% | 50% | 0% | 0% | 5% | 0% | 17% |
|  | NM = D = O | 0% | 33% | 33% | 0% | 33% | 0% | 6% | 3% | 0% | 13% |
|  | P | 0% | 0% | 0% | 100% | 0% | 0% | 0% | 0% | 35% | 0% |
|  | P = D | 0% | 50% | 0% | 50% | 0% | 0% | 8% | 0% | 26% | 0% |
|  | P = O | 0% | 0% | 0% | 50% | 50% | 0% | 0% | 0% | 26% | 17% |
|  | P = D = O | 0% | 33% | 0% | 33% | 33% | 0% | 6% | 0% | 22% | 13% |
|  | P = NM | 0% | 0% | 50% | 50% | 0% | 0% | 0% | 5% | 26% | 0% |
|  | P = NM = D | 0% | 33% | 33% | 33% | 0% | 0% | 6% | 3% | 22% | 0% |
|  | P = NM = O | 0% | 0% | 33% | 33% | 33% | 0% | 0% | 3% | 22% | 13% |
|  | P = NM = D = O | 0% | 25% | 25% | 25% | 25% | 0% | 4% | 3% | 19% | 11% |

1. The “current allocation” scenario, where the incremental budget allocation matches the existing allocation across cadres (i.e., the allocation proportions represent the relative total salary costs across the cadres in 2024). This allocation results in a uniform HRH expansion with staffing increase rates that are equal to HRH budget growth rate *R* for every cadre every year (See explanations in Appendix A.1.3).
2. The 31 “equal allocation” scenarios, where the incremental budget is allocated equally among each non-empty subset of the 5 cadres (including subsets of single cadres and the subset of all 5 cadres).
3. The “gap allocation” scenario, where the incremental budget is allocated proportionally to the estimated extra cost of each cadre that is expected to be needed to meet all services demand as simulated in the TLO model. See more details of preparing this scenario below Table 1.

Each scenario is simulated 10 times in the TLO model with an initial population size of 100,000 in 2010. The reported results are the mean and 95% confidence intervals of the outputs of ten runs, which are scaled to the true Malawian population size of 14.5 million in 2010. The HRH expansion process from 2025 to 2034 within the TLO simulation period from 2010 to 2034 is illustrated in Appendix A.1.2.

### Main analysis

First, we compare the performance of all scenarios in terms of health consequences, as measured by disability-adjusted life years (DALYs), and health service volumes, as measured by number of treatments delivered, in each making the comparison to the “current_allocation” scenario. Total DALYs and health service volumes are divided into 6 health areas: reproductive, maternal, newborn, and child health (RMNCH); HIV/AIDS; Malaria; TB; noncommunicable diseases (NCDs) and transport injuries. The specific causes of death or injuries in each health area are listed in Appendix A.1.6, which are among the leading causes of mortality and disability in Malawi (1, 17, 23, 24). In all result figures, the scenarios are ranked in the same ascending order of total DALYs.

Next, we conduct a two-step analysis to explore if there would be better allocation strategies than those defined above. In step one, we build a linear regression model (M1) to approximate the relationship between total DALYs incurred between 2025 and 2034, denoted by *y*, and average yearly increase rates of cadres, denoted by *x*_*C*_, *x*_*D*_, *x*_*NM*_, *x*_*P*_, *x*_*O*_; use Ordinary Least Squares method and the data of simulation results of 33 scenarios (data size = 330, with 10 simulations per scenario) to estimate the coefficients; and test the prediction performance of the regression model via another 100 randomly generated allocation scenarios by comparing the predicted and simulated outcome (data size = 100, with 1 simulation per scenario). In step two, we do an optimisation analysis (M2) to find the allocation strategy that minimises the DALYs, as approximated by M1. Here the average yearly increase rate of each cadre is constrained to the HRH budget growth rate, the incremental budget allocation proportion, the total salary cost of staffing of that cadre, and the total salary cost of staffing of all cadres in 2024 (See full constraint details in Appendices A.1.3 and A.1.4.). We then use this scenario in the TLO model in order to estimate the total DALYs and compare it to the others.

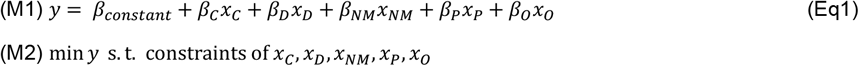

### Sensitivity analysis

The simulations in main analysis assumes a fixed level of incremental budget growth rate (i.e., *R* = 4.2%), perfect consumables availability (i.e., consumables are always available upon request) and default health system function (i.e., diagnostic accuracy, referral practice and HCW competence are informed by available data). We further conduct sensitivity checks by changing the incremental budget growth rate (i.e., *R* = 5.8%, *R* = 2.6%), setting default consumables availability (i.e., current availability level as informed by available data), or setting maximal health system function (i.e., perfect diagnostic accuracy, referral practice and HCW competence). See Appendix A.1.5 for full details of health system parameter assumptions made for the analyses and the resulting “gap_allocation” scenarios. Notice that the sensitivity checks focus on the relative performances of all 33 scenarios as included in Table 1 and do not include conducting optimal analysis.

## Data sources and availability

The data source for HRH staffing, patient-facing time and service time requirements is the Malawi Health Sector Strategic Plan (HSSP III) for 2023-2030 from the Ministry of Health, Government of Malawi (3, 9). The data source for HRH annual salary is Staff Returns from the Ministry of Health, Government of Malawi (21). All other data needed to run the TLO model simulation are available at this website: https://www.tlomodel.org/resources/index.html. Simulation outputs used to generate tables and illustrations are available at a public, open-access repository: https://doi.org/10.5281/zenodo.15792206.

### Patient and Public Involvement statement

Patients or the public were not involved in the design, or conduct, or reporting, or dissemination plans of our research.

## Data Availability

The data source for HRH staffing, patient-facing time and service time requirements is the Malawi Health Sector Strategic Plan (HSSP III) for 2023-2030 from the Ministry of Health, Government of Malawi. The data source for HRH annual salary is Staff Returns from the Ministry of Health, Government of Malawi. All other data needed to run the TLO model simulation are available at this website: https://www.tlomodel.org/resources/index.html. Simulation outputs used to generate tables and illustrations are available at a public, open-access repository: https://doi.org/10.5281/zenodo.15792206.

## Ethics statement

This research is approved by the College of Medicine Malawi Research Ethics Committee (COMREC, P.10/19/2820) in Malawi. Individual informed consent was not required as only anonymised secondary data was used.

## Transparency statement

The lead author affirms that the manuscript is an honest, accurate, and transparent account of the study being reported; that no important aspects of the study have been omitted; and that any discrepancies from the study as originally planned (and, if relevant, registered) have been explained.

## Results

### The health outcomes of alternative scenarios

Figure 1(a) shows the total DALYs averted in the period of 2025-2034 for each of the scenarios compared to the “current_allocation” scenario (See full numerical results in Appendix A.2.2). The nine scenarios on the left that simultaneously expand Clinical, Pharmacy and/or Nursing and Midwifery cadres would achieve greater health gains than the “current_allocation” scenario. The “gap_allocation” scenario generates the greatest health gains and averts DALYs by 3.86% (95% CI: [2.81%, 4.91%]), equal to 3.41 (95% CI: [2.46, 4.36]) million DALYs averted, compared to the “current allocation” scenario. The next-most impactful scenarios are the “C = P =NM” averting DALYs by 3.02% (95% CI: [2.04%, 4.00%]) and the “C = P” scenario with a comparable performance; and the other six scenarios have percent DALYs averted ranging between 0.75% and 2.28%.

**Figure 1.**
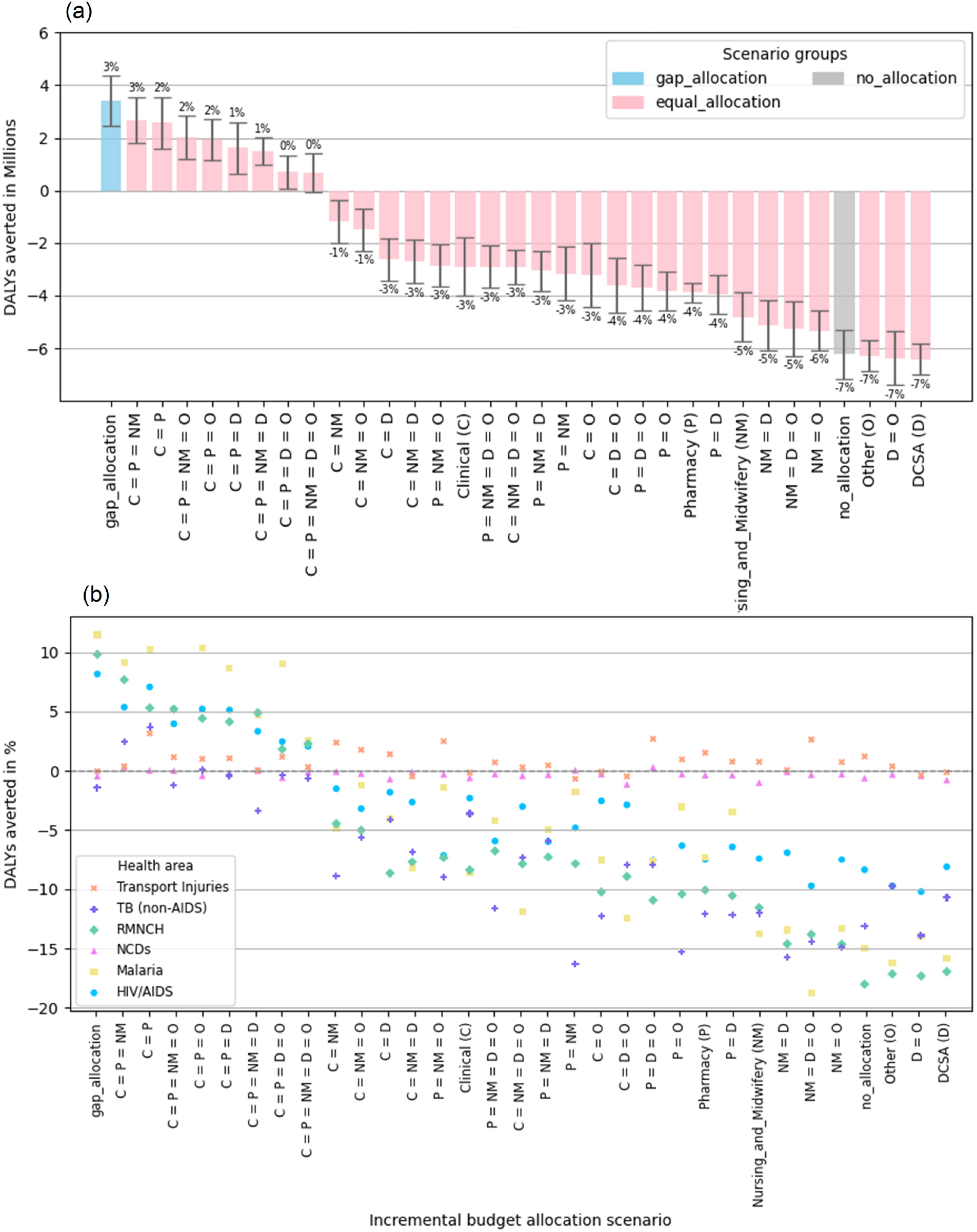
DALYs averted vs “current_allocation” strategy, 2025-2034. (a): Total DALYs averted (annotated with percentage changes), (b): Relative DALYs by cause area averted.

By contrast, all other scenarios would have worse health outcomes than the “current_allocation” scenario, with percent DALYs averted ranging between -7.30% and - 1.37% (where negative DALYs averted means more DALYs incurred). These include scenarios only expanding single cadres: for example, “Clinical (C)” -3.32% (95% CI: [-4.61%, -2.03%], “Pharmacy (P)” -4.43% (95% CI: [-4.89%,-3.96%]), “Nursing and Midwifery (NM)” - 5.49% (95% CI: [-6.58%, -4.40%].

Figure 1(b) shows the total DALYs averted split into health areas of RMNCH, HIV/AIDS, Malaria, TB, NCDs and Transport injuries (See Appendix A.2.1 for the figure with 95% CIs and Appendix A.2.2 for full numerical results). As compared to the “current_allocation” scenario, the nine better scenarios on the left that simultaneously expand Clinical, Pharmacy and/or Nursing and Midwifery cadres achieve significantly better health gains in the areas of RMNCH, Malaria and HIV/AIDS. Meanwhile, the other scenarios have worse health outcomes in the areas of RMNCH, Malaria, HIV/AIDS, and TB. The areas of NCDs and Transport injuries have no significantly different health outcomes across all scenarios by contrast.

### The health services delivered in alternative scenarios

Figures 2(a) and 2(b) show the absolute and relative increase of service volumes by health area across all scenarios as compared to “current_allocation” strategy (See the figure with 95% CIs in Appendix A.2.1 and full results in Appendix A.2.2). As expected, the overall patterns are consistent with those of the DALYs averted. The nine better allocation scenarios generally have more services delivered, by approximately up to 2% for Malaria, up to 3% for HIV/AIDS, RMNCH, TB and transport injuries, and up to 6% for NCDs; among which, the “gap_allocation” strategy would achieve the highest increases for these areas except Malaria.

**Figure 2.**
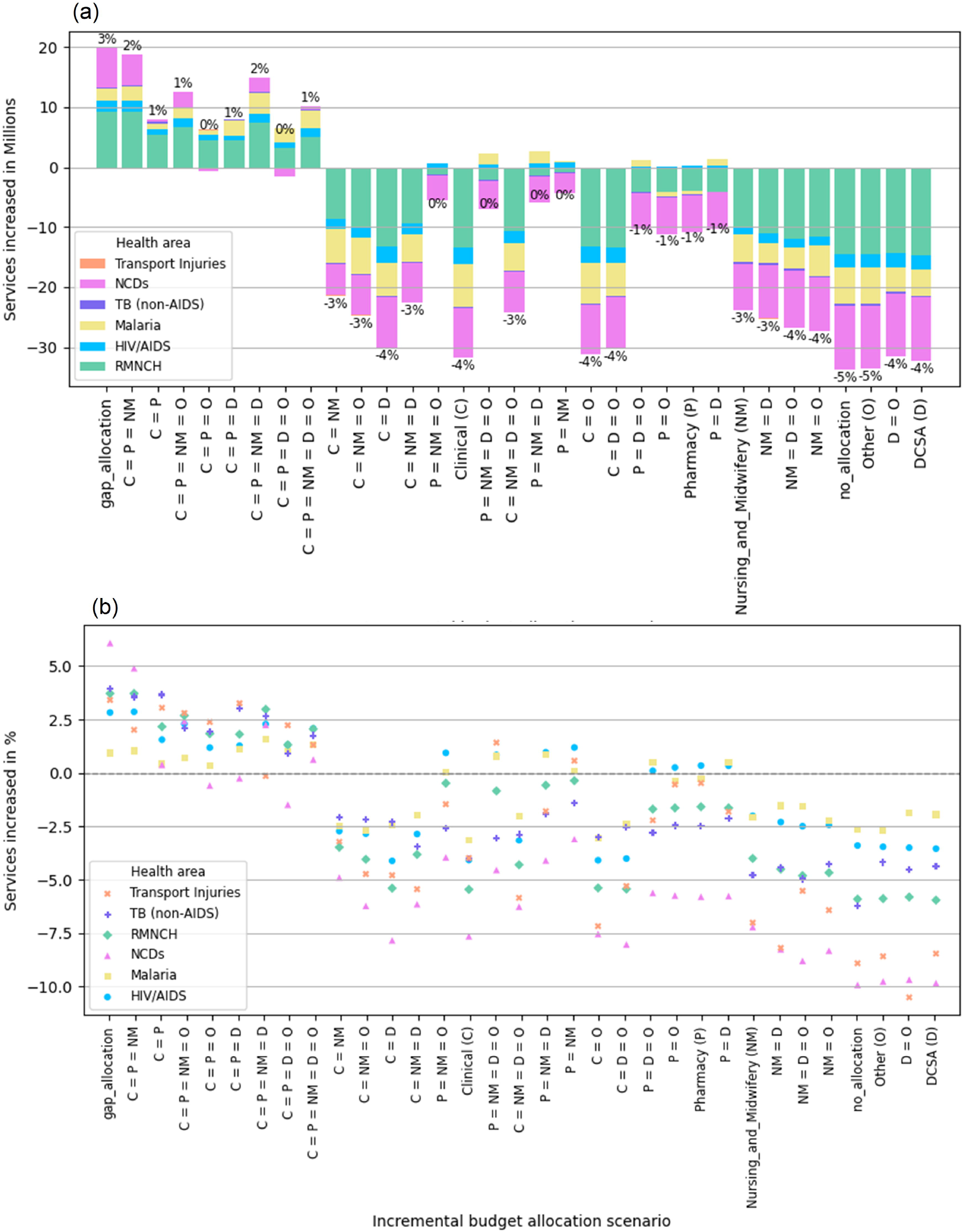
Increase in number of treatments by health area vs “current_allocation” strategy, 2025-2034. (a): Absolute service increase (annotated with total percentage changes), (b): Relative service increase.

On the contrary, the other worse scenarios would have fewer services delivered overall than the “current_allocation” scenario, and the worst scenarios could be equivalent to “no expansion”, which itself would have services decreased approximately by 3% for HIV/AIDS and Malaria areas, by 6% for RMNCH and TB areas, and by 9% for NCDs and transport injuries areas. Nonetheless, scenarios that expand Pharmacy cadre without Clinical cadre (e.g., “P = NM”) would have very modest decrease of total services and even significantly slight increases (< 1%) for Malaria and HIV/AIDS.

### Optimisation analysis

For step one, the estimated coefficients in regression model M1 are:

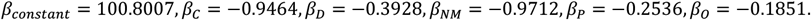

The adjusted R-squared is 0.801. To interpret the coefficients, taking *β*_1_ as an example, it indicates that a 1% average yearly increase of Clinical cadre staffing between 2025 and 2034 corresponds to a decrease of 0.9464 million DALYs in total in this period.

The prediction performance is tested using 100 randomly generated allocation scenarios. We find that 96% of the random scenarios have simulated DALYs within 95% prediction intervals. This shows good support of the regression model. More details can be found in Appendix A.2.3.

For step two, the “optimal solution” to optimisation model M2 that is detailed in Appendix A.2.4 is:

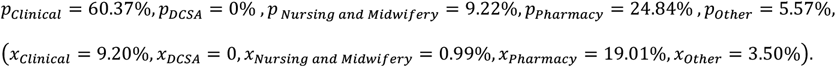

Running this scenario in TLO simulation shows that it would avert DALYs by 4.08% (95% CI: [3.34%, 4.83%]) as compared to the “current_allocation”, which is similar in magnitude to that achieved in the “gap_allocation”.

### Sensitivity analyses

As shown in Appendix A.3, the relative performances between in the 33 scenarios of “current”, “equal” and “gap” allocations are overall robust by changing incremental budget increase rate and decreasing consumable availabilities, or improving health system function. In particular, the “gap allocation” and “C = P = NM” scenarios remain the most impactful scenarios. Although, limited consumable availability would hinder the full potential of health benefits of different HRH investment strategies, with fewer DALYs would be averted as compared to “current allocation” scenario; and improvement of health system function would further shift the impacts of investing in different cadres, as Nursing and Midwifery cadre would have more impacts than Pharmacy cadre across the scenarios.

## Discussion

We find that HRH bottlenecks in achieving health gains are multifactorial: increasing capacity of one cadre has limited benefit, whereas using the same incremental budget to increase capacity of multiple cadres (i.e., Clinical, Nursing and Midwifery, and Pharmacy) would have a much greater benefit. Therefore, the health impact of HRH investment depends on the choice of how to allocate the budget between in cadres in a complex manner, and this has substantial implications for the health gains that may be achieved. The most impactful strategy we found was the “gap_allocation” strategy that aims to anticipate the needs for each cadre. We also found that a linear regression model could be estimated from the results of running a range of scenarios in the model, from which an estimate of the optimal allocation of resources among cadres could be found, which was equally impactful. However, these strategies may not have the same impact in practice as estimated here, due to unmodelled implementation constraints.

The relationship between capabilities of different HCW cadres and health outcomes is complex but was well represented by the linear model M1. This enabled us to develop a user-friendly tool (access [here](https://hrhexpansion-healthoutcome.streamlit.app/)) for predicting health consequences (as measured in total DALYs) of any incremental budget allocation strategy, for the purpose of supporting wider decision-making of HRH planning. In this tool, the user could input any proportion of the incremental budget allocated to each cadre (as long as they are non-negative and sum up to one), then it will translate the proportions into HRH increase rates and output the health consequences. This tool enables decision-makers to explore the approximate potential health outcome of any strategy of interest. However, the reliability is more limited than the full execution of the TLO model simulation.

The results in areas of NCDs and transport injuries show that changes in HRH capabilities and health service volumes do not correspond with changes in health outcomes, unlike areas of RMNCH, Malaria, HIV/AIDS, and TB. These are consistent with findings in previous research, suggesting that current service delivery scope as captured in the TLO model, rather than HRH capabilities, is constraining health gains achievement in these areas (1).

Overall, this research has presented a simulation-based method to understand the complex relationships between in HRH capability, health service provision and health outcome, and provided insights and tools that could inform relevant HRH policy making. Nonetheless, there are several limitations.

By focusing on expanding HRH capabilities in the near future under a limit annual incremental budget that is proportional to the total salary costs of current staffing, we did not consider the dynamics of current staffing, or take account of the costs of recruitment, training, supervision and retention for both current and expanded staff. We also assumed a fixed salary for each cadre in the simulation period. These other costs and potential salary increases can be easily embedded in the analysis once data becomes available.

We have assumed no task shifting/sharing among cadres, although it is happening in low-resource settings (25), as modelling it requires empirical evidence, Task shifting/sharing would change the health service provision and health outcomes of HRH expansion strategies, possibly in a good way because of more flexibility allowed; but might be in another direction if quality of care is not guaranteed (26). We have not considered potential changes in quality of care or HRH productivity brought by the expansion: the relationship between them remains an interesting research direction.

More flexibility of the HRH expansion process could be allowed, where the incremental budget allocation strategies can change every year or several years instead of being fixed throughout the period. Besides, the analysis is conducted on the national level: further analyses are required to inform policy making on granular levels of the health care system, such as community, primary, secondary and tertiary levels.

Other health system parameters such as health care seeking behaviour and prioritisation policies of service provision could also impact the health outcomes of HRH expansion strategies. In practice, we may expect care seeking behaviour or service prioritisation to change in response to the change of HRH capabilities.

Finally, the impact of DCSA and Other cadres including Laboratory, Mental and Radiography staff might be underestimated as their roles are the least well specified in the TLO model (3). The Ministry of Health in Malawi is planning to increase staffing of these cadres: further calibration of TLO model and exploration of health impacts of investing in these cadres are needed to inform such decision-making.

## Conclusion

Using the TLO model, we demonstrate that there is not a simple HRH bottleneck in achieving health gains in the context of complex interplay between demography, epidemiology, treatment scope and effectiveness, and health resource constraints in the health care system. As such, strategic HRH investment across multiple cadres can yield substantially greater health gains than uniform or single-cadre expansions; these decisions should ideally be informed by an anticipation of the healthcare needs that will be faced by the system in the years ahead. Overall, this analysis provides a practical framework for HRH planning in complex, resource-constrained settings.

