## Appendix for "Health impacts of expanding different health workforce cadres under a limited budget in Malawi"

### Appendices

#### A.1 Methods supplementary

##### *A.1.1 Current HRH count and salary*

Table A.1.1 HRH cadre

| **Cadre** | **Sub-cadre** |
|---|---|
| Clinical | Medical Officer /Specialist, Clinical Officer / Technician, Medical Assistant |
| DCSA | DCSA (Disease Control and Surveillance Assistant) |
| Nursing and Midwifery | Nurse Officer, Nurse Midwife Technician |
| Pharmacy | Pharmacist, Pharm Technician, Pharm Assistant |
| Laboratory* | Lab Officer, Lab Technician, Lab Assistant |
| Dental* | Dental Officer, Dental Therapist, Dental Assistant |
| Mental* | Mental Health Staff |
| Nutrition* | Nutrition Staff |
| Radiography* | Radiographer, Radiography Technician, Sonographer, Radiotherapy Technician |

The source for this table is (2). *These cadres are grouped into Other cadre in the main analysis.

Table A.1.2. HRH count, annual salary and salary cost per cadre.

| | Staff count | Annual salary (usd) | Salary cost (usd) |
|---|---|---|---|
| Clinical | 5,271 | 6,147 | 32,402,540 |
| DCSA | 14,804 | 2,361 | 34,951,557 |
| Nursing and Midwifery | 11,150 | 6,024 | 67,167,902 |
| Pharmacy | 789 | 5,069 | 4,002,083 |
| Dental Health* | 211 | 5,842 | 1,233,604 |
| Laboratory* | 1,101 | 5,769 | 6,354,070 |
| Mental Health* | 69 | 7,973 | 550,189 |
| Nutrition* | 91 | 4,770 | 434,478 |
| Radiography* | 269 | 6,352 | 1,709,537 |
| *Total* | 33,756 | NA | 148,805,961 |

*These cadres are grouped into Other cadre in the main analysis.

HRH count (as of 2024) is calculated from scaling up the 2019 data from the data source (1, 2) using yearly multipliers between 2020 and 2024. The yearly multipliers (2020: 1.027745, 2021: 1.076495, 2022: 1.076495, 2023: 1.076495, 2024: 1.076495) are calculated from Ministry of Health staff data as explained elsewhere (3). These multipliers are assumed uniform for all cadres due to unavailable data per cadre, thus the HRH data of 2024 may not accurately reflect the real staff distribution in 2024.

To derive the annual salary (as of 2018), we take the weighted average annual salary across salary grades (seniority) within each cadre, given the current distribution of salary grades in the workforce. Notice that the annual salary for Other cadre is the average annual salary among Dental health, Laboratory, Mental health, Nutrition and Radiography cadres.

##### *A.1.2 Illustrations of allocation scenarios and HRH expansion simulation*

Figure A.1.1 Illustration of any incremental budget allocation scenario.

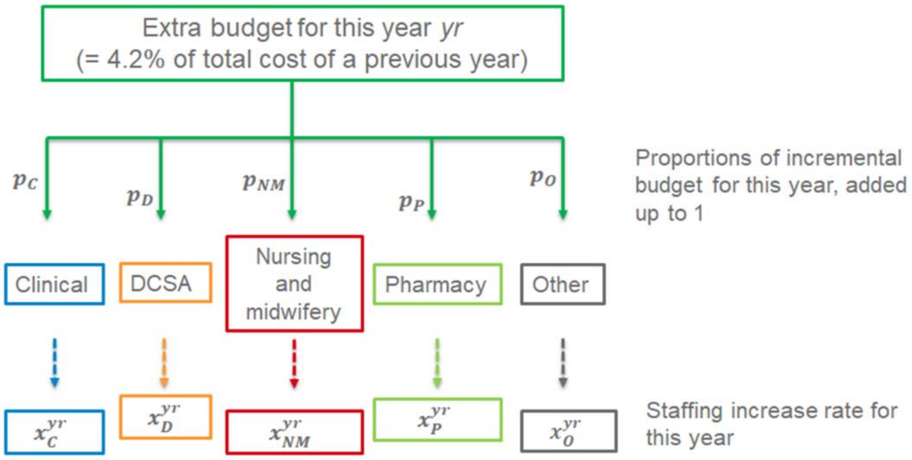


Figure A.1.2. The simulation and HRH expansion diagram.

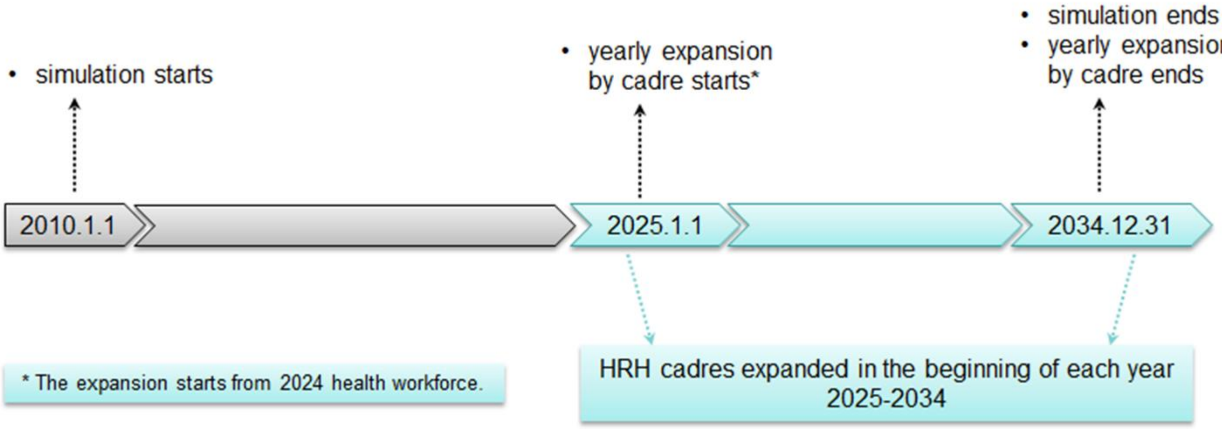


##### *A.1.3 Relationship between HRH increase rates and incremental budget allocation proportions*

Below are the equations to determine increase of each care in each year:

$$\textit{cadre}\text{ count of year }(yr+1) - \textit{cadre}\text{ count of year }yr$$

$$= \text{all cadres cost of year }yr \cdot R \cdot p_{cadre}/\textit{cadre}\text{ annual salary}$$

$$= \text{all cadres cost of year 2024} \cdot (1+R)^{yr-2024} \cdot R \cdot p_{cadre}/\textit{cadre}\text{ annual salary} \qquad \text{Eq(2)}$$

where $yr$ is each year from 2025 to 2034, $cadre$ is each one of the cadres in consideration, $cadre$ annual salary is fixed between 2025-2034, $R$ is the HRH budget growth rate, and $p_{cadre}$ is the proportion of incremental budget allocated.

From Eq(2) we can derive yearly staff count of each cadre

$$\textit{cadre}\text{ count of year }yr$$

$$= (\textit{cadre}\text{ count of year 2024}$$

$$+ \text{all cadres cost of year 2024} \cdot [(1+R)^{yr-2024} - 1] \cdot p_{cadre}/\textit{cadre}\text{ annual salary})$$

$$\textit{cadre}\text{ count of year 2034}$$

$$= \textit{cadre}\text{ count of year 2024} + \text{all cadres cost of year 2024} \cdot [(1+R)^{10} - 1] \cdot p_{cadre}/\textit{cadre}\text{ annual salary}$$

and yearly increase rate of each cadre

$$x_{cadre}^{yr+1}$$
$$= (cadre \text{ count of year } (yr+1) - cadre \text{ count of year } yr)/\text{cadre count of year } yr$$
$$= [(1+R)^{yr-2023} - (1+R)^{yr-2024}]/[cadre \text{ cost of year 2024}/(p_{cadre} \cdot \text{all cadres cost of year 2024}) + (1+R)^{yr-2024} - 1]$$

as well as the *average yearly increase rate* between 2025 and 2034

$$x_{cadre}$$
$$= (cadre \text{ count of year 2034}/cadre \text{ count of year 2024})^{1/10} - 1$$
$$= \{1 + p_{cadre} \cdot (\text{all cadres cost of year 2024}/cadre \text{ cost of year 2024}) \cdot [(1+R)^{10} - 1]\}^{1/10} - 1 \qquad \text{Eq(3)}$$

We see that $p_{cadre}$ and $x_{cadre}$ have a non-linear relationship and that the yearly increase rates $x_{cadre}^{yr+1}$ are not necessarily the same for each year, or equal to $x_{cadre}$. One exception is the "current_allocation" scenario, where $x_{cadre}^{yr+1} = x_{cadre} = R$, this is because $p_{cadre} = cadre \text{ cost of year 2024}/\text{all cadres cost of year 2024}$. Although we use average increase rate for simplicity in the analysis, the decision variable of the problem is still $p_{cadre}$ that would produce yearly increase rates over 2025-3034 that determine the HRH expansion in TLO model simulation.

In addition, we can also calculate total salary cost distribution across cadres in any year yr between 2025 and 2034 as below. The proportions of salary cost of a cadre within the total staffing cost are not necessarily the same in each year; unless $p_{cadre} = cadre \text{ cost of year 2024} / \text{all cadres cost of year 2024}$, for which the yearly salary cost proportions will always be equal to $cadre \text{ cost of year 2024 }/\text{all cadres cost of year 2024}$.

$$\text{Proportion of salary cost of } cadre \text{ in year } yr$$
$$= cadre \text{ count of year } yr \cdot cadre \text{ annual salary / total salary cost of all cadres of year yr}$$
$$= (\{cadre \text{ cost of year 2024} + \text{ all cadres cost of year 2024} \cdot [(1+R)^{Y-2024} - 1] \cdot p_{cadre}\} / [\text{all cadres cost of year 2024} \cdot (1+R)^{Y-2024}])$$

Finally, notice that in TLO model simulation, we need to divide $p_O$, the incremental budget proportion for Other cadre, to specific Dental health, Laboratory, Mental health, Nutrition and Radiography cadres. For each scenario, we do this division according to the current salary cost proportions within these cadres, making sure that they have the same yearly increase rates with each other that can be represented by $x_O$.

##### *A.1.4 The constraints of optimisation problem M2*

In this problem, the HRH expansion constraints are equivalent to $p_{cadre} \geq 0, \sum p_{cadre} = 1$, where $p_{cadre}$ can be replaced by a formulation of $x_{cadre}$ that is in reverse of Eq(3):

$$p_{cadre}$$
$$= [(1+x_{cadre})^{10} - 1] \cdot cadre \text{ cost of year 2024}/\{\text{all cadres cost of year 2024} \cdot [(1+R)^{10} - 1]\} \qquad \text{Eq(4)}$$

##### *A.1.5 Health system parameters and assumptions for main and sensitivity analyses*

Tables A.1.3 and A.1.4 specify the assumptions of main health system parameters in TLO model and the resulting "gap allocation" scenarios for main and sensitivity analyses. The incremental budget allocation proportions and HRH increase rates in the "gap allocation" scenarios are overall robust as health system parameters (in particular, budget growth rate and consumable availability) change.

Table A.1.3. Assumptions of health system parameters in TLO model.

| | Main analysis | Sensitivity analysis - more budget | Sensitivity analysis - less budget | Sensitivity analysis - default consumable availability | Sensitivity analysis - maximal health system function |
|---|---|---|---|---|---|
| HRH budget growth rate | 4.2% | 5.8% | 2.6% | 4.2% | 4.2% |
| Consumable availability | Perfect | Perfect | Perfect | Default | Perfect |
| Health system function | Default | Default | Default | Default | Maximal |
| Health care seeking | Default | Default | Default | Default | Default |
| Service prioritisation policy | Naive | Naive | Naive | Naive | Naive |
| HRH constraint mode | Rigid | Rigid | Rigid | Rigid | Rigid |
| Bed availability | Default | Default | Default | Default | Default |
| Equipment availability | Perfect | Perfect | Perfect | Perfect | Perfect |

Note the budget growth rates = 4.2%, 5.8% and 2.6% are assumed as equal to, greater than, and less than GDP growth rate following the existing study (4). With "Default", it means the values of the parameter is informed by available data(5). With "Naive", it means the services are prioritised in order of Childhood emergencies > Adult emergencies > Non-emergencies (6). With "Rigid", it means the health care worker cannot extend their normal working hours or shorten the treatment time per patient when the care demands exceed capabilities (6).

Table A.1.4. The "gap allocation" scenarios.

| Scenario group | Scenario labels | Incremental budget allocation proportions $p$ | | | | | Average yearly increase rates $x$ (of workforce size) | | | | |
|---|---|---|---|---|---|---|---|---|---|---|---|
| | | Clinical (C) | DCSA (D) | Nursing & Midwifery (NM) | Pharmacy (P) | Other (O) | Clinical (C) | DCSA (D) | Nursing & Midwifery (NM) | Pharmacy (P) | Other (O) |
| gap allocation | gap_allocation | 43% | 2% | 37% | 14% | 4% | 7% | 0% | 4% | 14% | 2% |
| | gap_allocation_more_budget | 43% | 2% | 37% | 14% | 4% | 7% | 0% | 4% | 14% | 2% |
| | gap_allocation_less_budget | 43% | 2% | 37% | 14% | 4% | 7% | 0% | 4% | 14% | 2% |
| | gap_allocation_default_cons | 43% | 2% | 38% | 14% | 3% | 7% | 1% | 4% | 14% | 2% |
| | gap_allocation_max_hs_func | 51% | 1% | 26% | 15% | 7% | 8% | 0% | 3% | 15% | 4% |

##### *A.1.6 Categorisation of causes of DALYs and services*

Table A.1.5. Categorisation of cause of DALYs in TLO model

| **Health area** | **Cause type** |
|---|---|
| HIV/AIDS | HIV/AIDS |
| Malaria | Malaria |
| NCDS | Bladder cancer, Breast cancer, Oesophageal cancer, Prostate cancer, Other adult cancers, COPD, Depression/Self-harm, Diabetes, Epilepsy, Heart Disease, Kidney disease, Lower Back Pain, Stroke |
| RMNCH | Childhood Diarrhoea, Congenital birth defects, Lower respiratory infections, Maternal Disorders, Measles, Neonatal Disorders, Schistosomias |
| TB | TB (non-AIDS) |
| Transport Injuries | Transport injuries |

Table A.1.6. Categorisation of treatment types in TLO model

| **Health area** | **Treatment type** |
|---|---|
| HIV/AIDS | HIV/AIDS |
| Malaria | Malaria |
| NCDS | Bladder cancer, Breast cancer, Oesophageal cancer, Prostate cancer, Other adult cancers, Cardio Metabolic Disorders, COPD, Depression, Epilepsy |
| RMNCH | Alri, Antenatal care, Contraception, Delivery care, Diarrhoea, Epi, Measles, Postnatal care, Schisto, Undernutrition |
| TB | TB |
| Transport Injuries | Transport injuries |

#### A.2 Main results supplementary

##### *A.2.1 Additional figures*

Figure A.2.1. Mean DALYs by health area averted (%) with 95% CIs vs "current_allocation" strategy, 2025-2034.

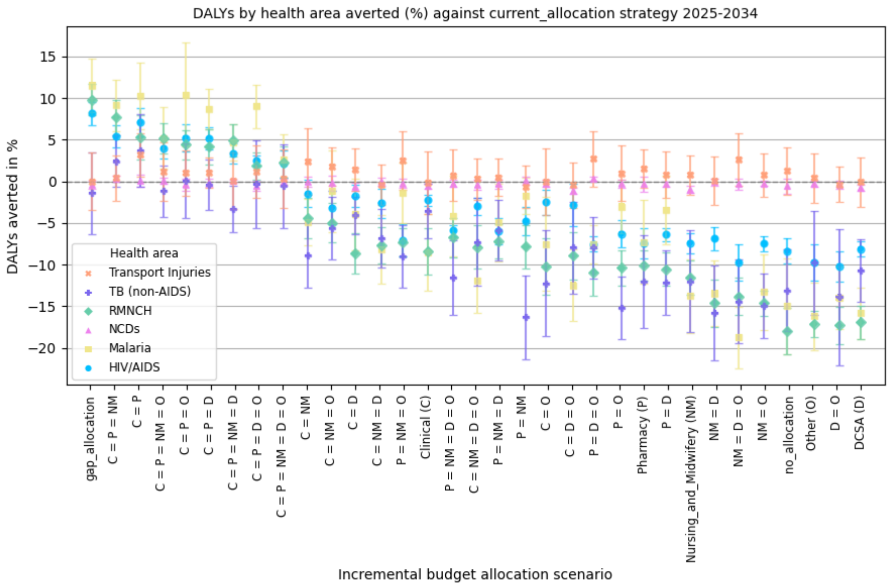


Figure A.2.2. Mean increase of service volumes (%) by health area with 95% CIs vs "current_allocation" strategy, 2025-2034

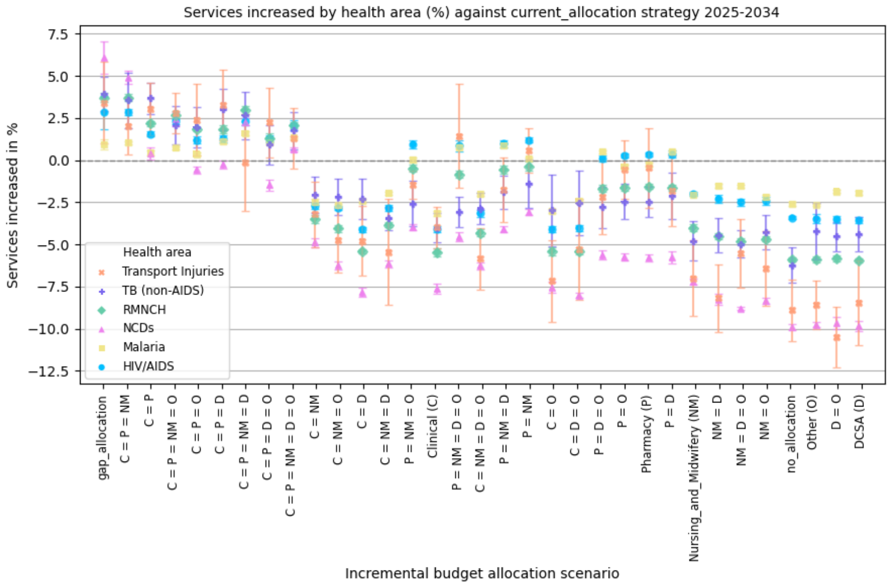


##### *A.2.2 Result datasets*

The full numeric results of number of DALYs and service volumes in main and sensitivity analyses can be found here: https://doi.org/10.5281/zenodo.15792206.

##### *A.2.3 Regression analyses*

###### *A.2.3.1 Performance of the regression model on HRH increase rates*

Figure A.2.3 (a) plots both the simulated and predicted DALYs of 33 allocation scenarios: the adjusted R-squared values is 0.801, indicating overall acceptable goodness of fit. Nevertheless, it tends to overestimate the DALYs of the better scenarios (including the "current_allocation") that simultaneously expand Clinical and Pharmacy cadres and underestimate the DALYs of the worse scenarios that expand Clinical or Pharmacy cadre without the other. We further test the overall prediction performance using 100 randomly generated allocation scenarios. Figure A.2.3 (b) shows the percentage of the random scenarios that have their simulated DALYs within the prediction intervals w.r.t different confidence levels of alpha, where the percentage would decrease as alpha increases. The prediction performance is confirmed acceptable as 96% of random scenarios have simulated DALYs within the 95% prediction intervals.

**Figure A.2.3**. Regression analysis of DALYs on HRH increase rate. (a): Fitted DALYs (with 95% Confidence Intervals and Prediction Intervals) vs Simulated DALYs, (b): Performance test by 100 random allocation scenarios.

(a)

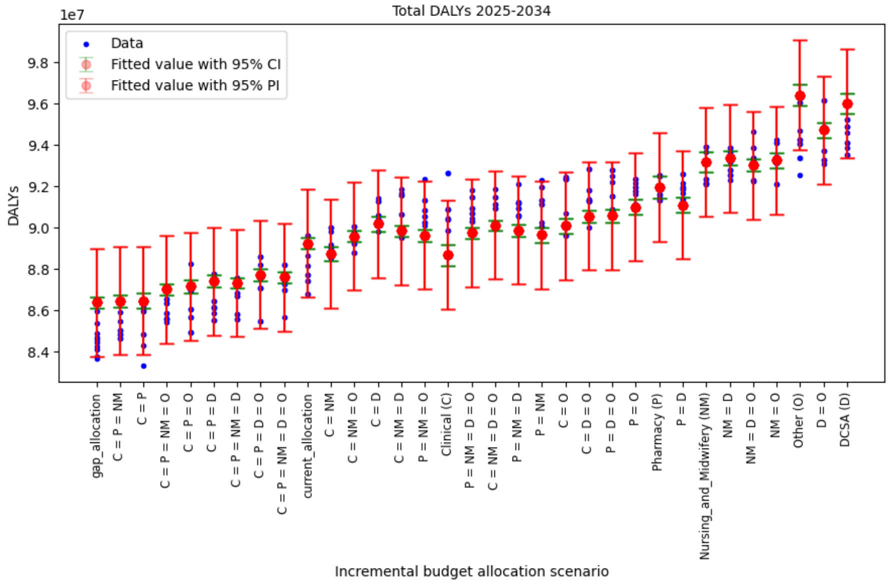


(b)

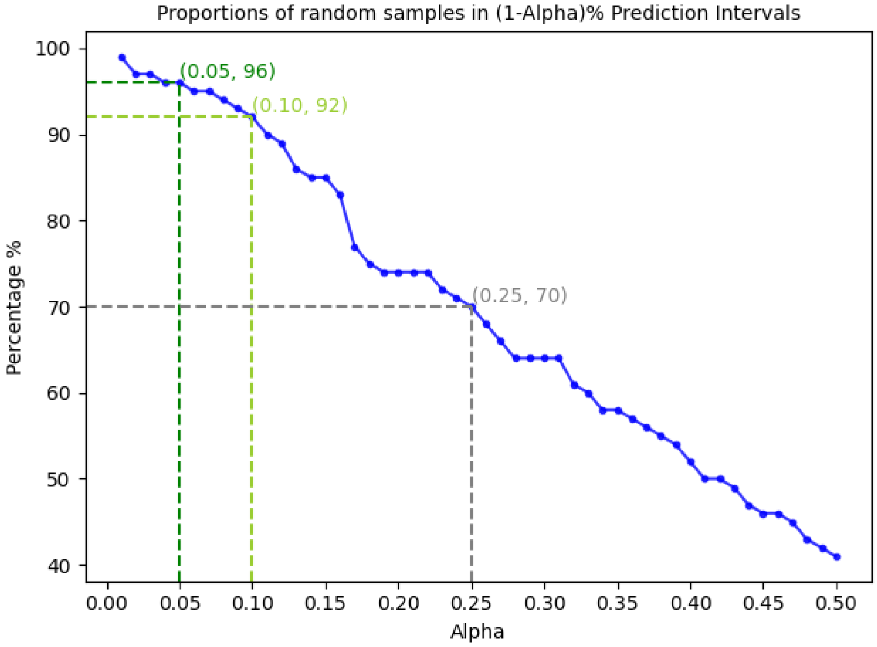

##### *A.2.3.2 Regression on incremental budget proportions*

We conduct linear regression analyses on incremental budget allocation proportions and on average increase rates separately. The model of the latter has much higher prediction accuracy than the former as indicated by the adjusted R-squared values (0.415 vs 0.801). Since average increase rates and incremental budget allocation proportions of cadre have a non-linear relationship as shown by Eq(2) and Eq(3), the two regression analyses have quite different performance on goodness of fit (see Figure A.2.3 above and Figure A.2.4 below).

Figure A.2.4. Regression analysis of DALYs incurred on incremental budget proportions, with adjusted R-squared value = 0.415.

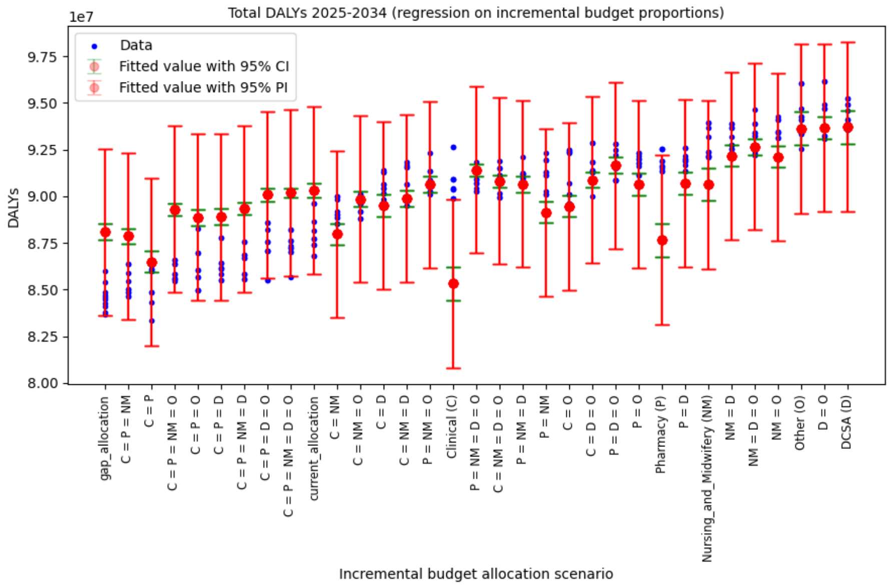


#### *A.2.4 The full optimisation problem M2*

Let $z_{cadre} = (1 + x_{cadre})^{10}$, then Eq(4) is transformed to

$$p_{cadre} = (z_{cadre} - 1) \cdot (cadre \text{ cost of year } 2024/\text{all cadres cost of year } 2024)/[(1+R)^{10} - 1].$$

Then, the constraints of $p_{cadre} \geq 0$ are transformed to $z_{cadre} \geq 1$, and the constraint of $\sum p_{cadre} = 1$ is transformed to $\sum z_{cadre} \cdot cadre \text{ cost of year } 2024/\text{all cadres cost of year } 2024 = (1+R)^{10}$.

By using all relevant data (M1 coefficient estimates, incremental budget growth rate, staffing and cost per cadre), the problem M2 is equivalent to

$$\text{M2}' \qquad \max\ -100.8007 + 100 \cdot [0.9464\left(z_{Clinical}{}^{\frac{1}{10}} - 1\right) + 0.3928\left(z_{DCSA}{}^{\frac{1}{10}} - 1\right) + 0.9712\left(z_{Nursing\ and\ Midwifery}{}^{\frac{1}{10}} - 1\right) + 0.2536\left(z_{Pharmacy}{}^{\frac{1}{10}} - 1\right) + 0.1851\left(z_{Other}{}^{\frac{1}{10}} - 1\right)]$$

$$\text{s.t. } z_{Clinical} \geq 1, z_{DCSA} \geq 1, z_{Nursing\ and\ Midwifery} \geq 1, z_{Pharmacy} \geq 1, z_{Other} \geq 1$$

$$0.2178\, z_{Clinical} + 0.2349\, z_{DCSA} + 0.4514\, z_{Nursing\ and\ Midwifery} + 0.0269\, z_{Pharmacy} + 0.0690\, z_{Other} = (1 + 0.042)^{10}$$

The corresponding feasibility problem is

$$\text{FP} \quad \text{s.t.} \quad 100.8007 - 100 \cdot [0.9464 \left(z_{Clinical}{}^{\frac{1}{10}} - 1\right) + 0.3928 \left(z_{DCSA}{}^{\frac{1}{10}} - 1\right) + 0.9712 \left(z_{Nursing\ and\ Midwifery}{}^{\frac{1}{10}} - 1\right) + 0.2536 \left(z_{Pharmacy}{}^{\frac{1}{10}} - 1\right) + 0.1851 \left(z_{Other}{}^{\frac{1}{10}} - 1\right)] \leq \text{a threshold of DALYs in million incurred}$$

$$z_{Clinical} \geq 1, z_{DCSA} \geq 1, z_{Nursing\ and\ Midwifery} \geq 1, z_{Pharmacy} \geq 1, z_{Other} \geq 1$$

$$0.2178\ z_{Clinical} + 0.2349\ z_{DCSA} + 0.4514\ z_{Nursing\ and\ Midwifery} + 0.0269\ z_{Pharmacy} + 0.0690\ z_{Other} = (1 + 0.042)^{10}$$

The problem M2 is a convex optimisation problem because the objective function to be maximised is s strictly concave function when the decision variables are not smaller than 1, as its Hessian matrix (as below) is negative-definite with all eigenvalues negative (as detailed below), and the constraints are always linear. This means that there exists an optimal solution. The software Lingo and package CVXPY in Python can solve the problem and give the optimal solution of increase rates. We then use Eq(4) to derive the corresponding optimal allocation proportions.

The Hessian matrix:

$$H = \begin{bmatrix} -8.5167 z_{Clinical}{}^{-1.9} & & & & \\ & -3.5352 z_{DCSA}{}^{-1.9} & & & \\ & & -8.7408 z_{Nursing\ and\ Midwifery}{}^{-1.9} & & \\ & & & -2.2824 z_{Pharmacy}{}^{-1.9} & \\ & & & & -1.6659 z_{Other}{}^{-1.9} \end{bmatrix}$$

and the eigenvalues are equal to the diagonal values of the Hessian matrix, which are negative as $z_{cadre} \geq 1$.

#### A.3 Sensitivity analyses

Here we summarise the findings from detailed results as presented in the subsequent subsections:

Changing the budget growth rate from 4.2% to 5.8% and 2.6% does not change the relative performance between in the scenarios: (1) The "gap_allocation" and "equal" allocation to Clinical and Pharmacy cadres (with and without Nursing and Midwifery cadres) rank top 3 and the better group of scenarios remains; meanwhile, more budget does bring more health benefits overall as compared to "no_allocation" but slightly restrains the relative advantages of the better scenarios as compared to "current_allocation". (2) The benefited areas from the investment would consistently be RMNCH, Malaria, TB and HIV/AIDS if to compare "current_allocation" and "no_allocation". However, the top 3 strategies do not have consistent advantages in TB area as compared to "current_allocation". (3) The better scenarios have more RMNCH, Malaria, TB and HIV/AIDS services delivered overall. (4) For NCDs and traffic injuries areas, significant increase and decrease of services would not correspond to significant change of health outcomes.

Changing the consumable availability from "perfect" to "default " (i.e., as informed by data ) does not bring much difference to the relative performance (in terms of DALYs and service volumes) between in the scenarios. But limited consumables availability does constrain the potential health benefits of HRH expansion, as the relative advantages of better scenarios to "no_allocation" and "current_allocation" both shrink.

Changing the health system function from “default” to “max” does change the relative performance of some scenarios. The “gap_allocation” and “equal” allocation to Clinical, Pharmacy and Nursing and Midwifery cadres remain the best and outperform the “current_allocation” in areas of RMNCH, HIV, TB and Malaria, with significant increase of services only in RMNCH and TB. However, scenarios that expand Clinical and Nursing and Midwifery cadres would have comparable or even better health outcomes (both in total and main areas) than those expanding Clinical and Pharmacy cadres without Nursing and Midwifery cadres, in particularly for RMNCH and HIV/AIDS. This indicates that the impact of Nursing and Midwifery cadre on health outcome is limited by health system functioning in terms of diagnostic accuracy, referral practice and HCW competence.

The regression models M1 all have acceptable adjusted R-squared values, although the prediction performances are not tested by random allocation scenarios as it was conducted in the main analysis.

##### *A.3.1 More budget*

DALYs of “no expansion” scenario: 93.90 (95% CI: [93.42, 94.38]) million. DALYs of “current_allocation” scenario: 85.83 (95% CI: [85.44, 86.21]) million. DALYs of “gap_allocation” scenario: 83.45 (95% CI: [83.02, 83.89]) million.

For regression model 1, the adjusted R-squared values is 0.781; the coeffficients are:

$$\beta_{constant} = 100.4043, \beta_C = -0.7492, \beta_D = -0.2993, \beta_{NM} = -0.7124, \beta_P = -0.2190, \beta_O = -0.1472.$$

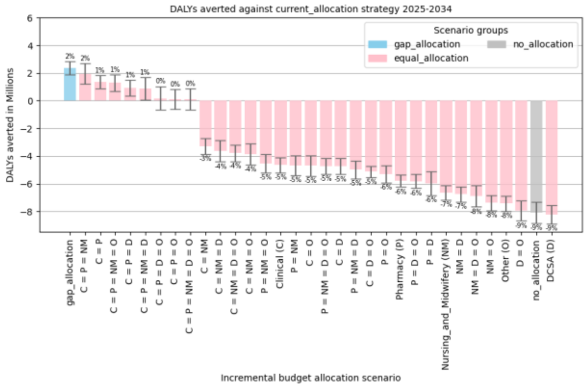


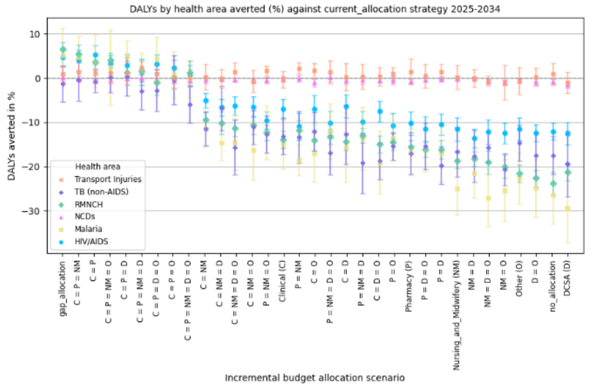


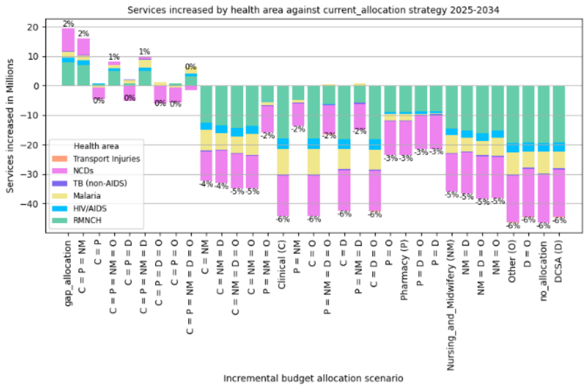


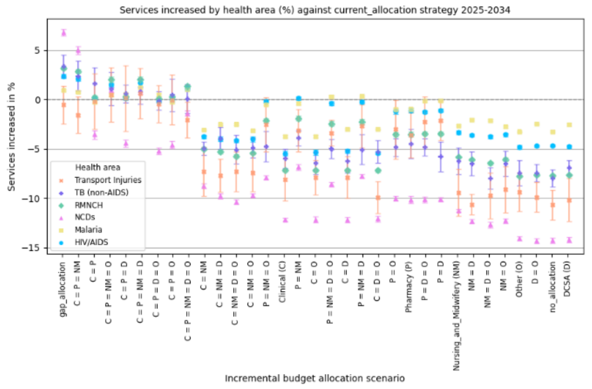

##### A.3.2 Less budget

DALYs of “no expansion” scenario: 95.45 (95% CI: [94.80, 96.11]) million. DALYs of “current_allocation” scenario: 90.84 (95% CI: [90.23, 91.45] million). DALYs of “gap_allocation” scenario: 87.21 (95% CI: [86.51, 87.90]) million.

For regression model 1, the adjusted R-squared values is 0.740; the coeffficients are:

$$\beta_{constant} = 102.3225, \beta_C = -1.3148, \beta_D = -0.6650, \beta_{NM} = -1.5711, \beta_P = -0.3221, \beta_O = -0.2774.$$

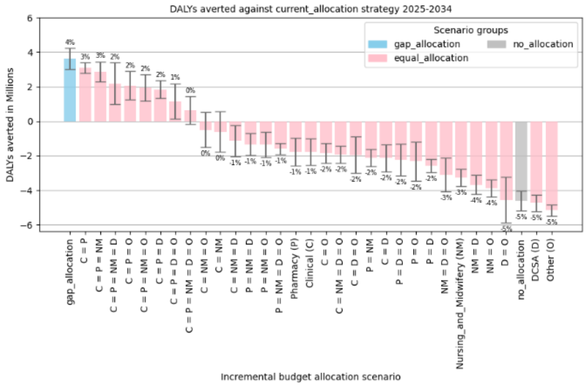


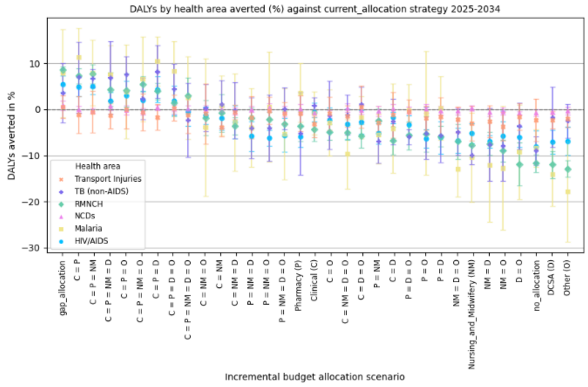


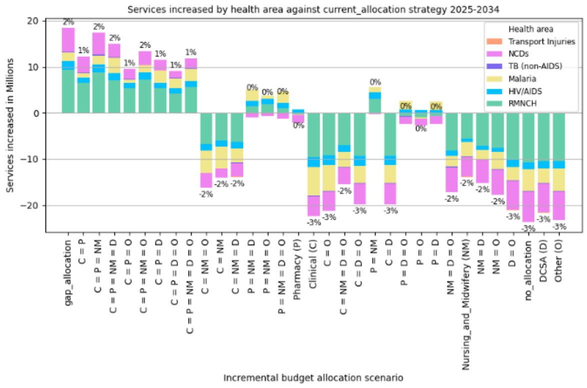


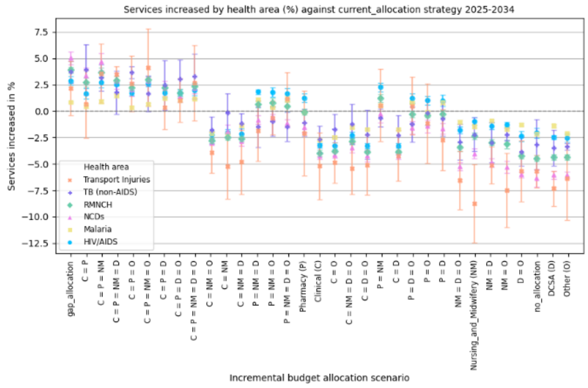


##### A.3.3 Default consumables availability

DALYs of “no expansion” scenario: 113.56 (95% CI: [113.00, 114.12]) million. DALYs of “current_allocation” scenario: 106.97 (95% CI: [105.70, 108.16] million). DALYs of “gap_allocation” scenario: 105.40 (95% CI: [104.56, 106.24]) million.

For regression model 1, the adjusted R-squared values is 0.645; the coeffficients are:

$$\beta_{constant} = 118.1100, \beta_C = -0.7652, \beta_D = -0.2997, \beta_{NM} = -0.8217, \beta_P = -0.1759, \beta_O = -0.350.$$

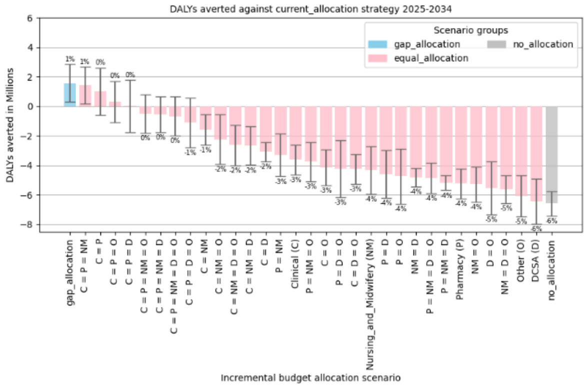


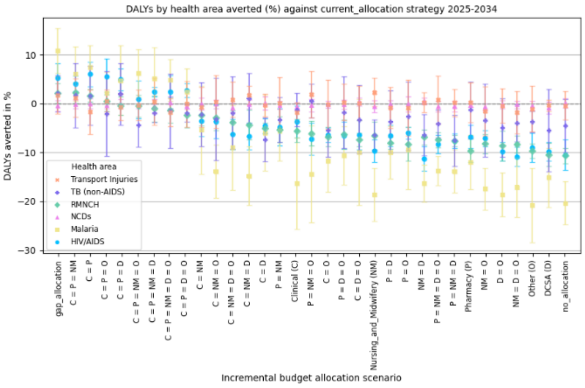

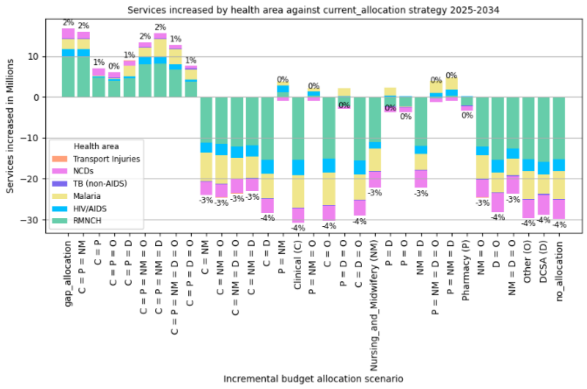


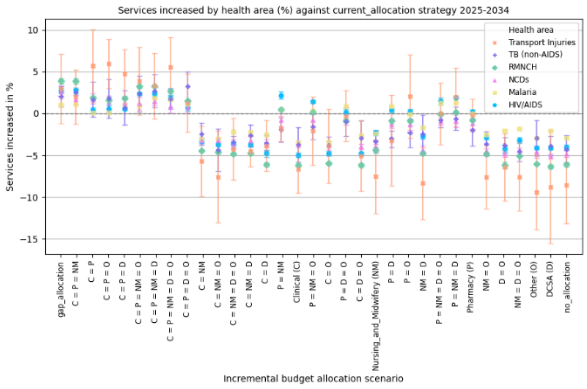


##### A.3.4 Maximal health system function

DALYs of “no expansion” scenario: 130.73 (95% CI: [129.21, 132.26]) million. DALYs of “current_allocation” scenario: 120.50 (95% CI: [119.18, 121.83] million). DALYs of “gap_allocation” scenario: 115.10 (95% CI: [114.18, 116.03]) million.

For regression model 1, the adjusted R-squared values is 0.768; the coeffficients are:

$$\beta_{constant} = 140.3744, \beta_C = -1.4983, \beta_D = -0.4446, \beta_{NM} = -2.0231, \beta_P = -0.220, \beta_O = -0.2145.$$

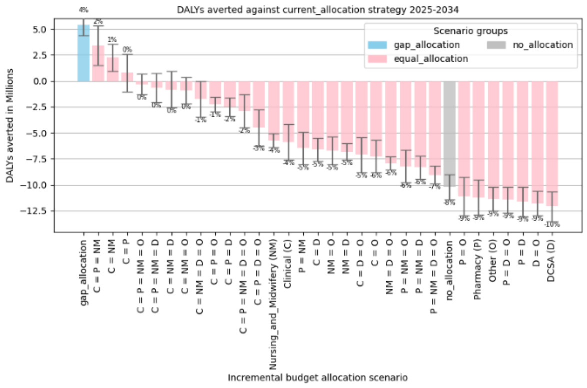


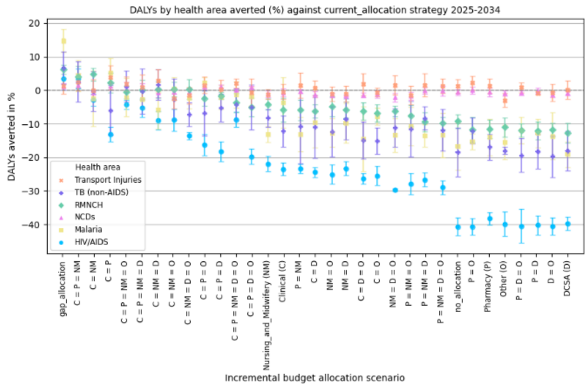


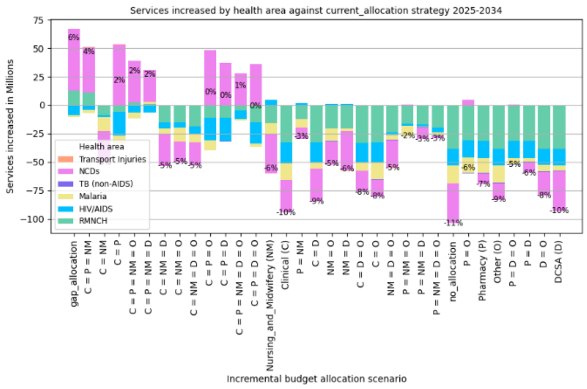


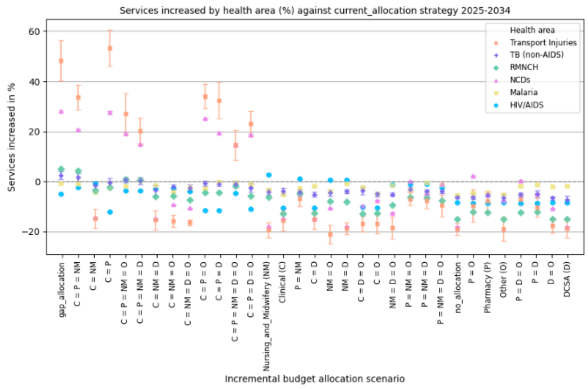
